# Is There a Health Inequality in Gambling Related Harms? A Systematic Review

**DOI:** 10.1101/19012104

**Authors:** J. N Raybould, M Larkin, R. J Tunney

## Abstract

**Objective:** Here we present a systematic review of the existing research into gambling harms, in order to determine whether there are differences in the presentation of these across demographic groups such as age, gender, culture, and socioeconomic status, or gambling behaviour categories such as risk severity and play frequency.

Primary and Secondary Outcome Measures: Inclusion criteria were: 1) focus on gambling harms; 2) focus on harms to the gambler rather than affected others; 3) discussion of specific listed harms and not just harms in general terms. Exclusion criteria were: 1) research of non-human subjects; 2) not written in English; 3) not an empirical study; 3) not available as a full article.

**Search:** We conducted a systematic search using the Web of Science and Scopus databases in February 2020. Assessment of quality took place using Standard Quality Assessment Criteria.

**Results:** 56 studies published between 1994 and 2019 met the inclusion criteria. These were categorised into thematic groups for comparison and discussion. There were replicated differences found in groups defined by age, socioeconomic status, education level, ethnicity and culture, risk severity, and gambling behaviours.

**Conclusion:** Harms appear to be dependent on specific social, demographic and environmental conditions that suggests there is a health inequality in gambling related harms. Further investigation is required to develop standardised measurement tools and to understand confounding variables and co-morbidities. With a robust understanding of harms distribution in the population, Primary Care Workers will be better equipped to identify those who are at risk, or who are showing signs of Gambling Disorder, and to target prevention and intervention programmes appropriately.

## INTRODUCTION

In their annual report of 2018 the Gambling Commission reported an overall increase of 1% in gambling participation of any form compared with data collected in 2017 (Gambling Commission, 2019c). The most popular form of gambling was found to be the National Lottery, and it was noted that there had been significant participation changes in several other areas. For example, there was an overall increase of 3.1% in sports betting between 2015 and 2018, and an increase of 1.9% in the use of fruit or slot machines. In the latest quarterly report there was an increase in areas such as arcades and lotteries, but also a large decrease of 10.3% in the profits of land based casinos (Gambling Commission, 2019b). Compared to this, remote online casinos had a 5.8% increase in profits. The changing face of gambling, and the introduction of remote gambling options, presents new challenges in understanding the nature of Gambling Disorder, and the harmful consequences associated with gambling behaviour. We predict that these harmful consequences are not distributed evenly amongst the population, and in conducting this review we aim to identify which individuals are most at risk, and how harms are likely to present in the general population before clinical diagnosis.

The fifth edition of the Diagnostic and Statistical Manual of Mental Disorders (DSM-5) (American Psychiatric Association, 2013) introduced Gambling Disorder as a behavioural addiction. This is the first behavioural addiction to be included in the DSM and the condition is an increasing public health concern (Gambling Commission, 2019a), as new accessible methods of play such as online and mobile gambling have led to an increase in new types of gambling behaviour. For diagnosis using the DSM-5, an individual must experience harmful consequences from their behaviour, so understanding the potential harms resulting from gambling is more important than ever.

There have been a number of recent Systematic Reviews completed in the field of gambling, investigating a range of ideas. For example, the relationship between crime and gambling disorders (Adolphe et al., 2019; Banks et al., 2019), quality of life measurement tools (Bonfils et al., 2019), comorbidity with other conditions (Marchetti, Verrocchio, & Porcelli, 2019), impulsivity in gambling (Ioannidis et al., 2019; R. S. C. Lee, Hoppenbrouwers, & Franken, 2019), or potential interventions and harm minimisation tools (Maynard et al., 2015; Pickering et al., 2018; Quilty et al., 2019; Tanner et al., 2017). Despite this body of research, and many individual studies investigating specific gambling harms, a systematic review of how harms are distributed across society hasn’t yet been done. Although many studies have investigated how harms can be minimised (Gainsbury, Abarbanel, & Blaszczynski, 2017; Harris, Parke, & Griffiths, 2018; Nerilee Hing et al., 2019), a complete understanding of the disease and its impact on society is dependent on understanding how harms are distributed across the population. If our intuitions are correct, then this poses a health inequality that needs addressing.

Harms related to gambling behaviour have been found to affect all types of individual, including low and moderate risk, or sub-clinical, gamblers (Browne & Rockloff, 2018; Rawat et al., 2017; Shaffer et al., 2004). The National Strategy to Reduce Gambling Harm (Gambling Commission, 2019d) states “An effective prevention plan must seek to identify the right mix of interventions to be applied at both the population and individual level,” and so a thorough understanding of how an individual experiences harm would be beneficial in understanding gambling as a whole and developing effective interventions. It has been suggested that in order to develop quality tools for minimising harm agencies need to consider aggregate harm to individuals, rather than the estimated prevalence of problem gamblers (Browne, Goodwin, & Rockloff, 2018). Current estimates suggest that there are 2 million adults experiencing some level of harm from gambling (Gambling Commission, 2019a) and a thorough understanding of how harms are presented within these individuals, and within at-risk groups, may help in identifying those at risk and targeting interventions where they are most needed.

### Objective

To present a systematic review of the existing research into gambling harms, to determine whether there are differences in the presentation of these across demographic groups such as age, gender, culture, and socioeconomic status, or gambling behaviour categories such as risk severity and play frequency.

## METHOD

### Search Strategy

In conducting the review we followed the Preferred Reporting Items for Systematic Reviews and Meta-Analysis (PRISMA) which can be seen in The PRISMA Checklist (Additional File 1). Studies that have explored specific harms and the prevalence of these within a population were identified using a search of records held by Web of Science. The database was searched on 25^th^ February 2020 using the following criteria; *(TI= ((gambl^*^ OR problem^*^ gambl^*^) AND (harm^*^ OR negative impact^*^ OR adverse impact^*^ OR detrimental impact^*^ OR negative ?ffect OR adverse ?ffect OR detrimental ?ffect OR consequences)))*. This yielded 195 results, which can be seen in detail in the Full Search Report (Additional File 2). An initial search within abstracts yielded 2,283 results, and the first two pages of these were not relevant to the review, so the search was restricted to titles only. Search terms were chosen using ‘thesaurus.com’ (Thesaurus.com) and the Oxford English Dictionary Online (Oxford University Press) to identify synonyms and the final criteria was developed through a trial and error process, using different synonym and Boolean term combinations until the most successful criteria were identified, in terms of number and relevance of studies returned. The criteria was then adapted to search records held by Scopus. This search yielded only a single result and so both titles and abstracts needed to be included. This second run yielded 134 results, giving a combined 329 studies from both websites.

### Inclusion Criteria

Studies were included that discussed harms from gambling, discussed harms to the gambler, and discussed or listed a minimum of two identified harms rather than discussing harm minimisation or harms in general terms. For the purpose of this review a clinical diagnosis of Gambling Disorder, or an increased risk of diagnosis as shown by the Problem Gambling Severity Index (PGSI) (J. Ferris & Wynne, 2013) or South Oaks Gambling Screen (SOGS) (Lesieur & Blume, 1987), was considered for discussion. Although an increased risk of diagnosis using the PGSI or SOGS cannot be considered an actual gambling harm, as not all at risk gamblers progress to develop Gambling Disorder, increasing scores on these measures do correlate with harms experienced (Angus et al., 2019; Holtgraves, 2009).

### Exclusion Criteria

We screened the 329 results to identify duplicates, studies unrelated to gambling, and those that did not discuss harms. We also excluded results not available in English, those that were articles, letters or books, studies that only covered the notion of harm without giving examples, those that only investigated harms to others, or harms from other related sources, and studies that were not available as a full article. This left 56 studies for review of which 19 were qualitative, 35 were quantitative, and 2 were of mixed methods design.

### Quality Evaluation

Studies were assessed for quality by two researchers using the Standard Quality Assessment Criteria (Kmet, Lee, & Cook, 2004). This measures the quality of both quantitative and qualitative research using a series of standardised questions, and we evaluated studies that followed a mixed methods approach in terms of the most prominent research style. The results of this assessment can be seen in the Table of Quality Checks (Additional File 3). Studies were coded in Excel using the guidelines set out by Kmet, Lee and Cook and coloured using a traffic light system for reviewing. Disagreements of more than one degree were discussed to reach a consensus, and scores were then combined to find an average.

### Data Analysis Plan

We extracted study design, participant number, diagnostic measures, harms measures, groups analysed, and funding source from the studies and identified categories for analysis. Full extracted data can be seen in the Table of Extracted Data (Additional File 4). We divided studies into these categories for comparison, with several studies providing results for multiple groups, and then summarised the results in brief.

For extracting relevant comparable data, we have used the 73-Item checklist developed by (2016)Langham et al. which identifies 8 domains of harm (Table 1). Delfabbro and King (2019) argue that certain items attributed as harms are labelled incorrectly; they suggest that chasing losses, gambling to obtain more excitement, or betting above affordable means, are behaviours that lead to harm and not the harms themselves. Schellinck et al. (2015) also argue that borrowing money is not a harm, but is in fact a predictor for the harms, debt and relationship conflict. Critical appraisal of the defined harms used in each study is therefore necessary.

**Table 1.** Langham et al. (2016) Taxonomy of Harm Domains

| Domain | Items Include but are not Limited to... |
| --- | --- |
| Financial Harms | Erosion of Savings<br>Bankruptcy |
| Relationship Disruption, Conflict or Breakdown | Dishonest communication<br>Social Isolation |
| Emotional or Psychological Distress | Distorted cognitions or erroneous beliefs<br>Extreme distress |
| Decrements to Health | Reduced self-care<br>Ongoing disability |
| Cultural Harm | Reduced engagement in cultural rituals<br>Extreme cultural shame |
| Reduced Performance at Work or Study | Tiredness and Distraction<br>Loss of Job |
| Criminal Activity | Vulnerability to illegal activities<br>Arrest and/or conviction |
| Life-course and Intergenerational Harms | Loss of life-course event i.e. marriage<br>Homelessness |
| A sample of harms identified by Langham, Thorne (Langham et al., 2016) |  |

### Patient and Public Involvement

There was no involvement from the general population or any individual with a Substance Addiction or Behavioural Addiction Disorder in this systematic review.

## RESULTS

### Search and Selection Results

The database searches returned 329 papers for review and 7 of these were excluded as duplicates. Analysis of titles and abstracts led to a further 59 exclusions for not discussing gambling, 67 exclusions for not discussing harms, or for discussing harms from other related sources such as gambling adverts, and 3 for investigating non-human test subjects. The remaining 193 studies were reviewed in full, resulting in 37 studies being excluded because they were articles, book chapters, letters or editorial pieces that discussed the topic without providing new information. Studies discussing only harms to others resulted in 13 exclusions, 4 studies were unobtainable, and 11 studies were not available in English. During the full-text review studies previously thought to discuss harms were removed for not discussing specifics (32), or discussing harm minimisation (40) rather than harm analysis, leaving 56 studies remaining.

**Figure 1.**
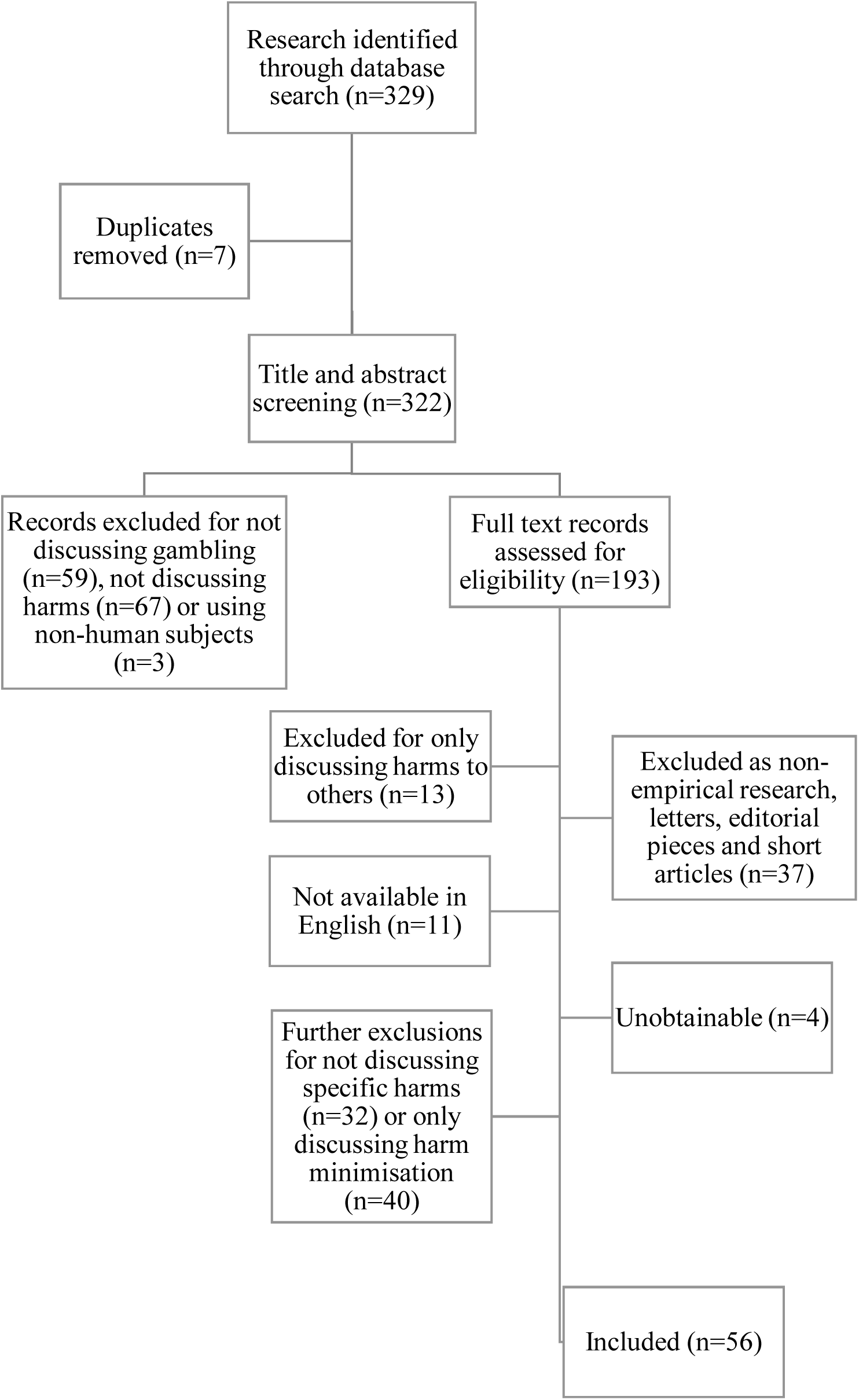
Preferred Reporting Items for Systematic Reviews and Meta-Analysis (PRISMA) Flowchart of Exclusions (Moher et al., 2009)

## MAIN RESULTS

### Description of Included Studies

Of the 56 studies included in this review, 14 used interviews, 2 used observation, 4 used focus groups, 26 involved surveys and 10 conducted reviews. Of the 10 reviews, 7 were systematic, and the remainder were narrative. Secondary data analysis was conducted in 14 of the studies, 2 studies used case notes, 1 study used online forum analysis, and 1 used health state valuation vignettes.

The most common funding bodies for this selection of studies were the Ministry of Social Affairs and Health Helsinki (5) and the Victorian Responsible Gambling Foundation (6). In total, the government funded 11 studies, medical institutes and foundations funded 3, gambling focused institutes or trusts funded 17, Colleges and Universities funded 5, research councils and institutes funded 3, and Star City Casino in Sydney funded 1. Of the remaining 16 studies, 11 received no funding and 5 did not declare their funding status.

**Table 2.**
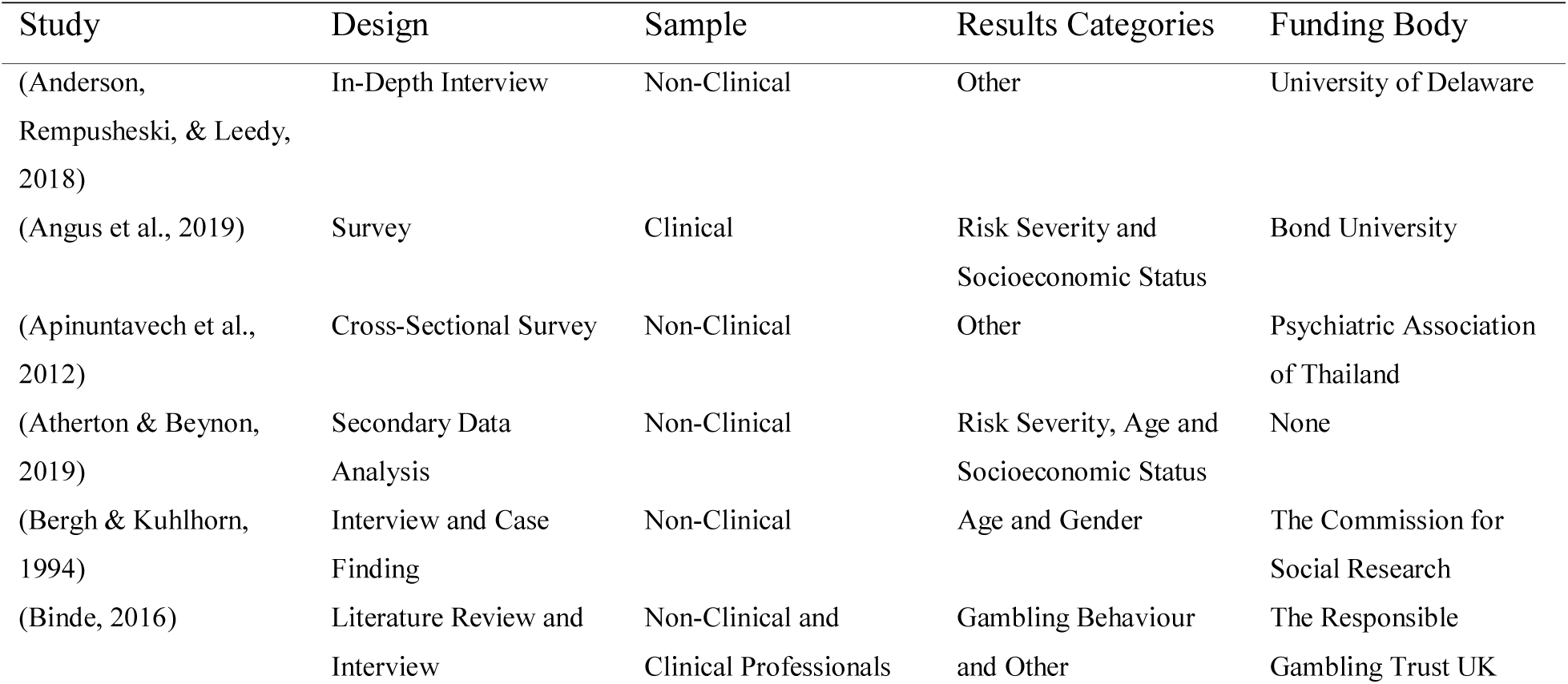
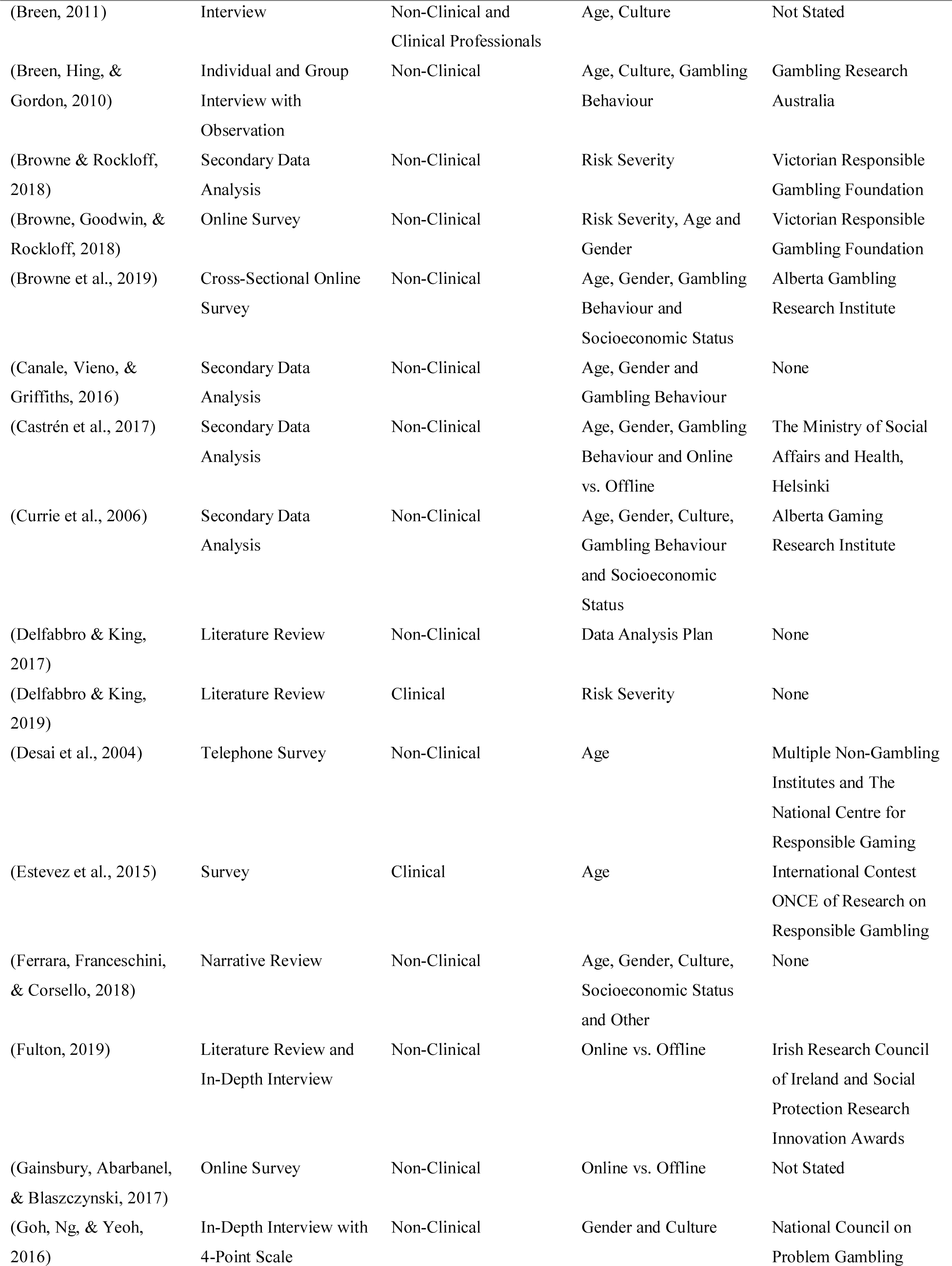
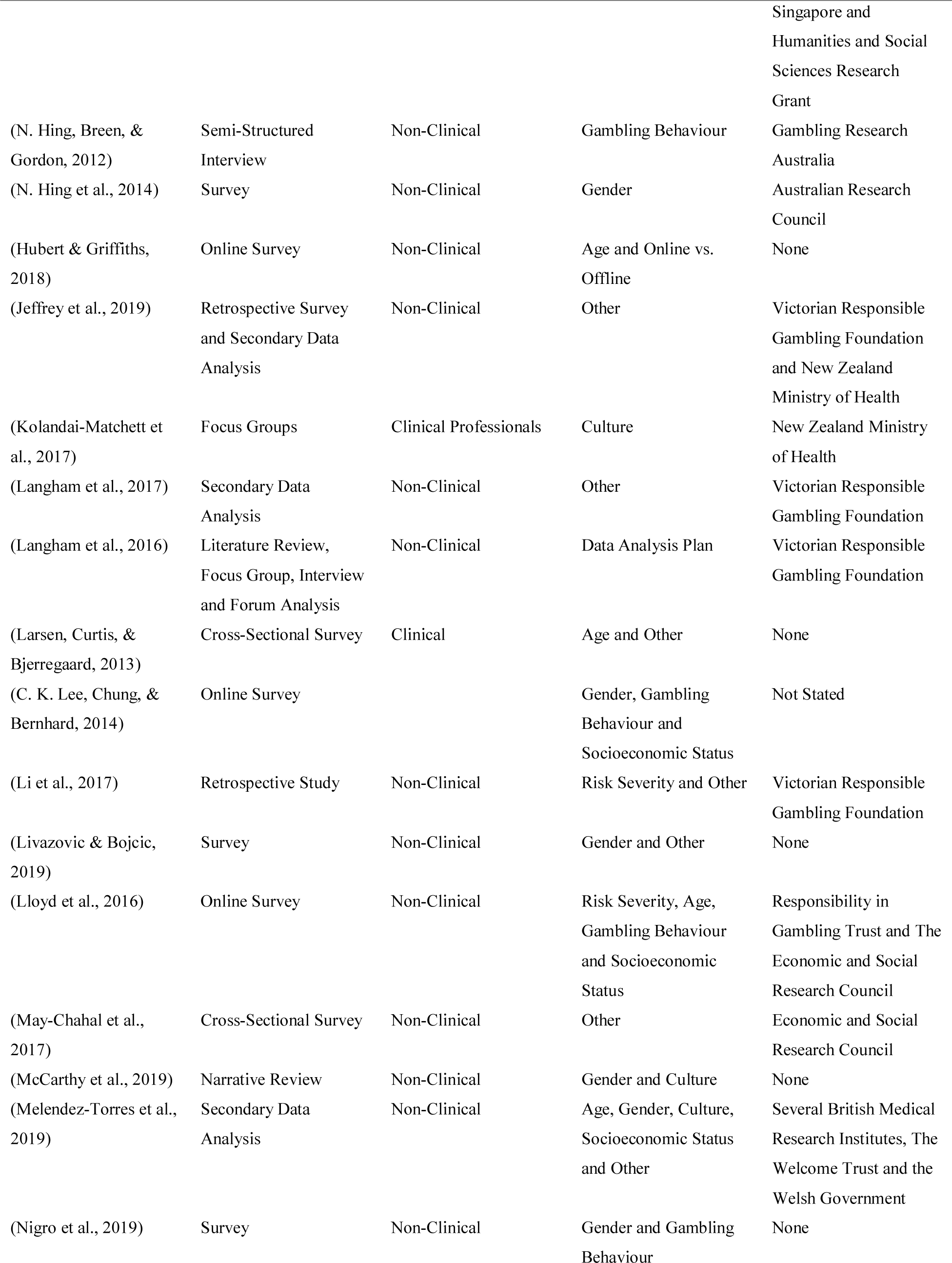
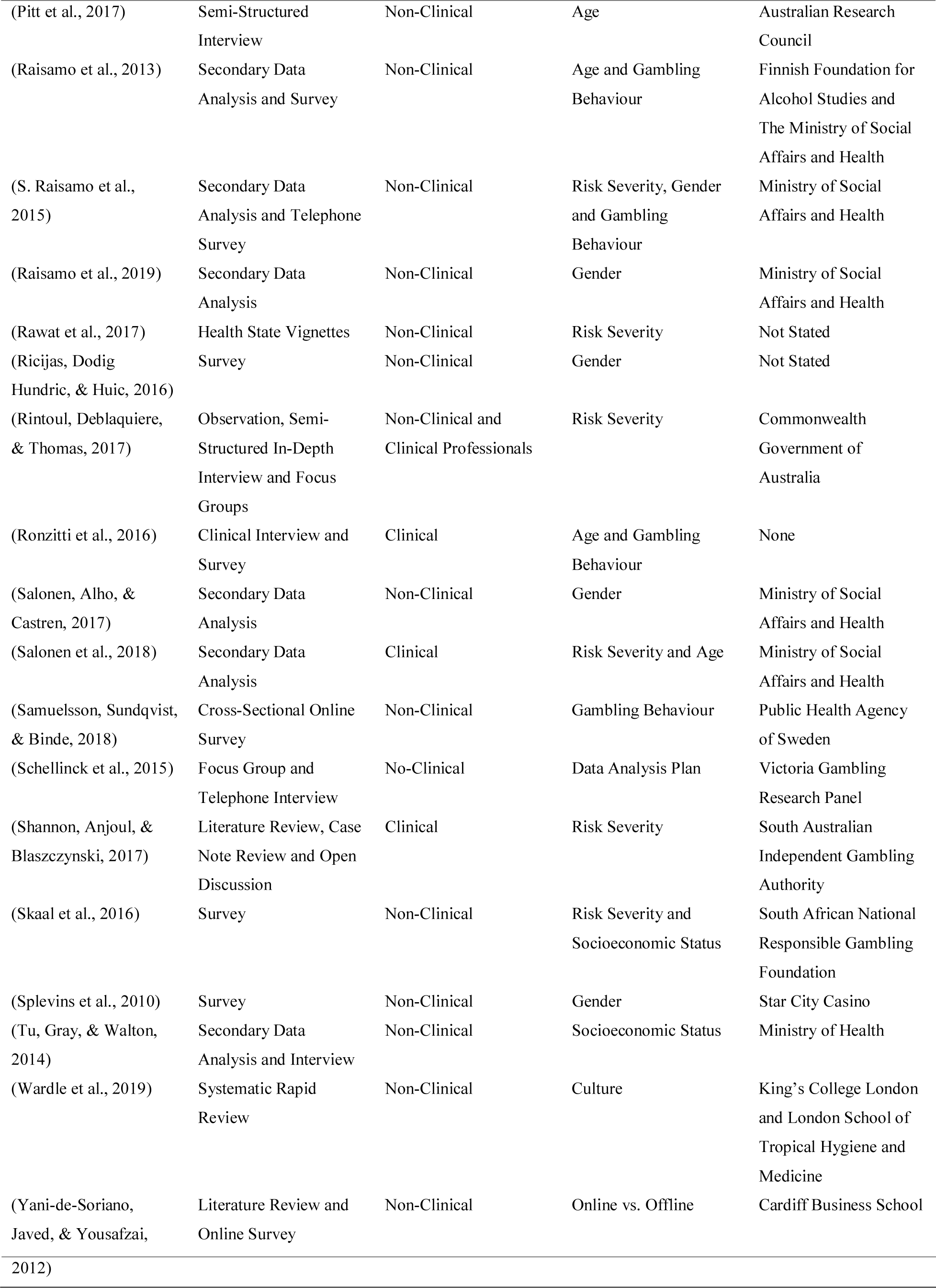
Summary of Data Extracted

### Risk Severity

Fourteen studies include data on risk severity, which is the measure of behaviour that puts someone at risk of developing a problem with gambling or developing Gambling Disorder. Rintoul, Deblaquiere, and Thomas (2017) found that venues looked for signs of problem gambling behaviour such as ‘shouting at the machine or other people in the venue’, appearing depressed, being withdrawn or emotional, excessive sweating, extended play and continuing to gamble with the winnings. Although the venue considered these items to be gambling harms it could be argued that continuing to gamble with the winnings is a predictor of financial harm rather than a harm itself. In 34 hours they observed these signs in all venues and concluded that risk factors for the behaviour included multiple machine use at one time and withdrawing money several times at the venue.

In academic research the risk of developing Gambling Disorder is most frequently measured using the PGSI, a component of the Canadian Problem Gambling Index (J. W. Ferris, H., 2001), which categorises an individual as low risk or non-problem (0 score), moderate risk (1-2 score), high risk (3-7 score), or a problem gambler (8+ score). These scores increase the confidence of accurate diagnosis but do not provide any clarity on the nature or severity of harms experienced by the individual. It is therefore important to state again that risk severity cannot be considered a gambling harm, as high-risk scores do not always lead to actual experienced harms or diagnosis. However, Shannon, Anjoul, and Blaszczynski (2017) found that the PGSI reliably correlated with 48 indicators of harm. These harm indicators were compiled using data from a scoping literature review, clinical case notes in a treatment setting, and a cohort of seven specialist clinicians.

Atherton and Beynon (2019) found that people who self-reported as a problem gambler were twice as likely to see their general practitioner for support with mental health concerns, five time as likely to be hospital inpatients, and eight times as likely to seek counselling. Angus et al. (2019) also found that the number of harms experienced increased with PGSI classification, and significantly less non-problem, low risk, or moderate risk gamblers reported harms compared with problem gamblers. Problem gamblers were also more likely to come from the clinical sample, who had significantly greater severity of harms in all domains, and of which 100% reported psychological harms, compared to only 54.69% in the community.

Many of the studies looking into risk severity focused on gathering evidence of the Prevention Paradox. This describes a situation where the majority of cases of a disease come from a population at low or moderate risk of that disease. In the case of gambling this means, the majority of harms are found within the low to moderate risk gamblers, and the minority is found within high risk or problem gamblers.

In one study the prevalence of harms across non-problem (PGSI), or non-pathological (SOGS), gamblers were twice that of problem gamblers in their participant sample (Browne, Goodwin, & Rockloff, 2018). Browne and Rockloff (2018) found that only 10% of financial harms across the study population were in the problem gambling or pathological gambling groups and that more than 50% of cases where someone sold their belongings to fund their gambling were in recreational or low risk gamblers. In contrast to this, they found that more than 50% of social deviance harms are found within problem gamblers, and the remaining categories of harm were evenly distributed across the severity groups.

Evidence of the prevention paradox was found by several other studies (Raisamo et al., 2015; Rawat et al., 2017; Salonen et al., 2018), though it was also found that the highest severity harms were generally only present in the problem gambling groups, as defined by the PGSI. Skaal et al. (2016) found psychological distress was only associated with problem gambling and Lloyd et al. (2016) found that self-harm thoughts were associated with problem gambling. Rawat et al. (2017) found that problem gamblers show similar disability weights to those of Bipolar Disorder or alcohol dependence, whereas the low risk group show disability weights equal to moderate anxiety. Disability weights are a health-related measure of quality of life using a ratio scale between 0 and 1, representing ideal health and death. These estimates further suggest that although the majority of harms may be found in the lower risk groups, the more severe harms are only prevalent within problem gambling groups.

Li et al. (2017) found that selling personal items, absence from work or study, reduced performance, poor sleep and extreme distress had the highest correlation with PGSI categories. They also found that reduced spending on essential expenses, absence from work or study, feelings of worthlessness, increased relationship conflict, and feeling like an outcast were the most effective discriminators between the low and high-risk groups.

Shannon, Anjoul, and Blaszczynski (2017) found that across both a clinical and non-clinical sample the lowest reported harms were substance use, suicide, bankruptcy, and education problems. They found that the distribution of averaged harm scores was consistent across both samples, excluding reduced savings and decreased happiness, suggesting that psychological and financial harms are the most significant. The most common gambling harms found represented negative impacts on general quality of life and psychological well-being; however, the mean level of harm for the 15 most common indicators in the community sample was less than one empirically defined unit.

Despite these results, there has been some criticism of the PGSI as a measure of gambling behaviour. Victorian Responsible Gambling Foundation (2019) categorises the PGSI groups as non-problem gambling, low risk, moderate risk and problem gambling, which is distinctly different to the categories used in some of these studies. Delfabbro and King (2019) argued that harms cannot be confidently attributed to gambling in low risk individuals, as some studies consider those who score a 0 on the PGSI as low risk, rather than no risk. They also argue that harms are not appropriately scaled, citing that “suicide is not equal to shame,” and that when controlling for severity low risk categories show very few harms. However, they did find that in financial harm categories the items they considered as more severe, such as selling belongings, were present even in the low risk groups. This may be due to the less affluent socioeconomic status of the low risk category, and it is hard to attribute these financial harms to gambling.

### Age

Twenty-one studies include data on age, and several of these found that being younger was associated with a higher risk of experiencing gambling harms (Atherton & Beynon, 2019; Browne et al., 2019; Castrén et al., 2017; Currie et al., 2006; Desai et al., 2004; Ferrara, Franceschini, & Corsello, 2018; Raisamo et al., 2013; Ronzitti et al., 2016). In particular one study found that younger age groups (16-34) were at risk of dependence and social harms Canale, Vieno, and Griffiths (2016), and Ferrara, Franceschini, and Corsello (2018) found that as well as higher rates of what they label “problematic gambling”, younger age groups showed a higher comorbidity with other addictions. Desai et al. (2004) found that younger gamblers were more likely to abuse alcohol and other substances, be incarcerated, and be bankrupt and depressed, and older gamblers were more likely to report good subjective general health.

In contrast, Melendez-Torres et al. (2019) found that harms experienced increased with age; however, they only researched participants attending school who would be categorised in the younger age groups of other studies. Some studies found that younger gamblers were less at risk of financial harms (Pitt et al., 2017; Raisamo et al., 2013), despite one suggesting that they spent more (Breen, Hing, & Gordon, 2010). And Larsen, Curtis, and Bjerregaard (2013) found that alcohol use increased with age in lifetime problem gamblers, as defined by the DSM-IV criteria for ‘pathological gambling’, in opposition to the trend seen in a general population.

Two studies found that age had no impact on harm profiles (Browne, Goodwin, & Rockloff, 2018; Salonen et al., 2018), and Lloyd et al. (2016) found no association between age and gambling-induced thoughts of self-harm. Pitt et al. (2017) found that children aged 8-16 showed little or no current harms as they were gambling at home with their families, spending small amounts of pocket money, or betting with activities such as push-ups against family members.

Despite the apparent absence of harms in the youngest age groups children developed false beliefs around gambling, such as that skill can be used to win, or that it is necessary for everyone to try gambling at least once. Children were also found to understand how gambling could gain them money. In Breen (2011) it was found that youth who were exposed to gambling at a young age were more likely to gamble later in life to increase their income, and that youth who missed school had reduced lifelong aspirations and reduced opportunities.

Further research is needed to understand the distribution of harms across age groups as it was found by Estevez et al. (2015) that sensation seeking and impulsivity were high in young gamblers. Anxiety, depression and psychoticism were partially mediated by impulsivity, and somatisation, obsessive-compulsive behaviour, interpersonal sensitivity, paranoid ideation and hostility were perfectly mediated. Hubert and Griffiths (2018) found that online gamblers are more likely to be younger, and Bergh and Kuhlhorn (1994) found that individuals aged 20-34 gambled for almost twice as long per session compared to participants over 35. Breen, Hing, and Gordon (2010) discovered that younger gamblers preferred to play poker or other card games, while Ferrara, Franceschini, and Corsello (2018) found that younger gamblers were more likely to be sports bettors. These additional variables could explain some of the variance in harm presentation and help to target appropriate interventions, as Ferrara, Franceschini, and Corsello (2018) found that 0.2-12.3% of adolescents in their study met the criteria for a diagnosis of Gambling Disorder depending upon country of residence.

### Gender

Twenty studies examined gender, and only a few of these found evidence that harms affected men and women differently (Bergh & Kuhlhorn, 1994; Browne, Goodwin, & Rockloff, 2018; Browne et al., 2019; Castrén et al., 2017). Where researchers found differences between genders, these were explainable by other factors. Though some studies show men have a higher prevalence of harms than women, (Currie et al., 2006; Ferrara, Franceschini, & Corsello, 2018; Nigro et al., 2019; Ricijas, Dodig Hundric, & Huic, 2016), we know that on average men gamble more frequently and spend more money when gambling (Canale, Vieno, & Griffiths, 2016; C. K. Lee, Chung, & Bernhard, 2014; Melendez-Torres et al., 2019; Raisamo et al., 2015). N. Hing et al. (2014) found that females from small villages and men from towns were more likely to be heavy commercial gamblers, however the harms suffered were the same and so this was likely due to usage level rather than gender.

Livazovic and Bojcic (2019) found that males in Croatia scored significantly higher on psychological, social, and financial consequences than females. However, they also scored significantly higher on risk behaviour and were more likely to score as a problem gambler on the Canadian Adolescent Gambling Inventory. Raisamo et al. (2015) found that although the prevalence of harms was higher in males, when controlling for frequency of play and amount spent gender was no longer a significant factor. Moreover, Splevins et al. (2010) found that men started gambling earlier than women did and found it more exciting. This led to increased spending and therefore an increased risk of harms such as substance use and interpersonal conflicts.

Despite this some studies have suggested key differences in how gambling harms present between genders. In Singapore, Goh, Ng, and Yeoh (2016) reported that “tentative evidence. points to the risk of child neglect when the problem gambler is the mother.” They also found that verbal abuse was most commonly males towards their mother, but found no difference in cases of physical abuse between genders. McCarthy et al. (2019) found that women were more likely to report mental health comorbidity than males, however causality is not discussed, and Raisamo et al. (2019) found that although the most common harm was guilt for both genders, the second most common was disrupted schoolwork for females and conflict with friends for males.

One key study that found an opposing result was Salonen, Alho, and Castrén (2017) who found that while gambling was more common in young males, women displayed an increase in specific harms between 2011 and 2015 where men did not. This particular study also looked at attitudes towards gambling and found that while female attitudes were generally negative over the age of 25, male attitudes were generally positive for all but the 15-17 age range. As the study used self-report data these differences in attitude could have affected how individuals reported harms.

### Culture

Eleven studies include data on culture and the majority of these discuss Australia and New Zealand. The included studies largely focus on single groups or comparing indigenous people and migrants to a society, so there are currently significant gaps that future studies may address. Although direct comparison is therefore not possible, some estimates and inferences can be made.

Kolandai-Matchett et al. (2017) found that Pacific New Zealand people experienced gambling through collectivist cultural values, meaning that additional harm dimensions were present. Some of the listed cultural harms include a loss of belonging or isolation, shame; loss of the community’s respect; disruption of trusting relationships; transference of communal responsibilities; and an overall loss of social cohesion. In a quotation from one of the interviewed participants, the researchers noted that the wider collective might exclude non-present or non-contributing members of the society.

Breen (2011) studied indigenous Australians and found that one key harm was the neglect of children when a parent gambles, and the eldest daughter would become their main caregiver. They also found that many people would gamble within a group, increasing their behaviour, but they would feel shame from losses and from the potential gossip within their close community, and Breen, Hing, and Gordon (2010) also found that a large concern for Indigenous Australians was the spread of a harms impact throughout the community. The range of harms shown within indigenous Australian communities included gambling away their pensions, relationship issues (Breen, Hing, & Gordon, 2010), betting above their means, guilt and shame, chasing losses (Breen, Hing, & Gordon, 2010), and to a lesser extent financial harms like borrowing money or selling items, and health problems. However, it is important to consider whether betting above their means, chasing losses, and borrowing or selling can be considered harms or simply predictors of harm.

McCarthy et al. (2019) suggested that women from ethnic minorities, indigenous communities and specifically Maori and Pacific women in New Zealand were more vulnerable to gambling harms than European women were. Melendez-Torres et al. (2019) also found that participants from white ethnicities were less likely to feel guilt from gambling, and a non-white British background was associated with more harms. Ferrara, Franceschini, and Corsello (2018) found that non-white males were most at risk of developing a gambling problem and, in the UK, Wardle et al. (2019) found that although migrants were less likely to gamble they were more likely to experience harms than individuals born in the country. By contrast, Currie et al. (2006) found that white men were more likely to report harms, and are more likely to gamble overall, with a higher frequency and higher spends.

The final study that investigated culture is that of Goh, Ng, and Yeoh (2016) who found that families in Singapore were at risk of acute financial harms when the problem gambler was a parent. Most households suffered double financial harms through loss of income and large debt. When the gambler was a mother without income, they found that the father would leave employment to care for the children, resulting in an income reduction for the entire household. Goh, Ng, and Yeoh (2016) also found that many people in Singapore viewed gamblers as self-centred, and siblings would often give up on them, rather than accepting problem gambling as an illness.

### Gambling Behaviour

One of the most predictable influences on gambling harms is the specific gambling habits and behaviours of the individual. Fourteen studies include data on gambling behaviours and several studies have agreed that a higher frequency of play, and higher amount of spending per session, leads to more harms (Canale, Vieno, & Griffiths, 2016; Castrén et al., 2017; Currie et al., 2006; Raisamo et al., 2015; Raisamo et al., 2013; Samuelsson, Sundqvist, & Binde, 2018). In particular, Nigro et al. (2019) reported that higher involvement in gambling was associated with higher levels of depression and self-reported memory impairment. N. Hing, Breen, and Gordon (2012) also reported that heavy gambling led to participants spending their entire pay or pension, borrowing money, and playing all night and day in both commercial and card games. Once again we need to consider whether borrowing money can be considered a harm, however in partnership with spending their entire pension it is clear that some financial mismanagement is present.

Game choice also affected harms, as heavy commercial gamblers reported more harms than card players did. These included debts, relationship issues, loss of home, missed bills, no food and low nutrition, child neglect and abuse from stress, lying, domestic violence, depression, suicidality, criminality, and selling their belongings.

Some other studies have found links between the quantity of harms and the choice of game type. Castrén et al. (2017) found that six out of twelve game type predictors were associated with more harmful consequences, including scratch games, betting, slot machines, non-poker online games, online poker, and non-monopoly games. They found that lottery play caused the lowest number of harms, and this finding is consistent with findings reported by Currie et al. (2006) who found that frequency of play on lottery games did not increase the harms experienced, whereas electronic gambling machines, ticket gambling, bingo and casino games did. Ronzitti et al. (2016) also concluded that Fixed Odds Betting Terminals and casino tables were associated with the highest scores on the PGSI, and gambling machines showed high PGSI scores compared to other methods of play, despite similar play times.

Two studies within the search looked at motivations for gambling, and although Browne et al. (2019) found no link between motivation of play and harms, C. K. Lee, Chung, and Bernhard (2014) found that excitement, escape and challenge motives were linked with positive outcomes, but financial motivation led to harms. Lloyd et al. (2016) also found that self-harm thoughts were associated with money as a motivator but was negatively associated with enjoyment motivations. Within this category it is also important to discuss factors such as player skill level and whether gambling is taking place in a group or individually, however none of the studies included in this review examined these specific areas.

### Online vs. Offline Gambling

As well as specific game type five studies look at the broader categories of online or offline gambling. Castrén et al. (2017) found only a weak link between online gambling and an increase in harms, however Gainsbury, Abarbanel, and Blaszczynski (2017) found that online gamblers tended to have higher PGSI scores. Moreover, online gamblers showed more variation in their choice of game and gambling behaviours, and in comparison to offline gamblers, they showed no preference for skill-based games.

Yani-de-Soriano, Javed, and Yousafzai (2012) found that online gambling was associated with binge drinking, cigarette smoking and an increased risk of developing problem gambling. However, they did not find a link between what they labelled as internet addiction and gambling. Hubert and Griffiths (2018) also found a link between online gambling and alcohol dependence, and they discovered that online gamblers were less likely to have jobs, children and a stable relationship, leading to unemployment and less money later in life. They further found that online gamblers were less able to control impulsivity and frustration, but despite this, they had fewer suicidal thoughts than offline gamblers, although actual suicide attempts were comparable in both groups.

Feelings of shame appeared to be lower in online gamblers relative to offline gamblers (Fulton, 2019), and it was suggested that online gamblers feel less judged since their behaviours could be more secretive and private. Despite these reduced feelings of shame, Fulton (2019) observed that secretive gambling increased financial harms due to the likelihood of concealed debt; and by living a double life secretive gamblers experienced increased stress, relationship conflicts, and emotional deterioration.

### Socioeconomic Status

There were eleven studies examining the socioeconomic factors that influenced the presentation of gambling harms, and all but one study concluded that less affluent socioeconomic groups are more at risk of experiencing harms than more affluent groups. Melendez-Torres et al. (2019) found that households that are more affluent were associated with more gambling behaviour and subsequently more harms. In comparison Ferrara, Franceschini, and Corsello (2018) found that individuals in routine socioeconomic groups were at the highest risk of developing an addiction to gambling, and Currie et al. (2006) concluded that participants who reported harms were more likely to be in a lower income bracket, and to have received no further education than high school. Similarly, Atherton and Beynon (2019) found that lower income households spent a higher proportion of their income on gambling, and were more likely to bet more than they could afford to lose, and Angus et al. (2019) found that clinical participants had significantly lower incomes than a community sample and a higher proportion of them reported harms.

Skaal et al. (2016) reported that urban residents were more likely to report psychological distress and be at a high risk of problem gambling on the PGSI. In comparison, harms were associated with employment status in peri-urban areas, with unemployment doubling the risk of problem gambling. Browne et al. (2019) found that income and education level are indirect risk factors of harm, and Lloyd et al. (2016) found that gambling related thoughts of self-harm were more prevalent in the unemployed.

C. K. Lee, Chung, and Bernhard (2014) reported that married people with a low income are more likely to be financially motivated to gamble, and this motivation could be associated with greater harms. Lloyd et al. (2016), however, found no link between problem gambling and marital status.

In an apparent contrast with other results, Tu, Gray, and Walton (2014) found that people in managerial or professional occupations appeared more likely to participate in gambling than people in routine occupations. However, although gambling rates in the most affluent groups dropped during times of recession, the rates within deprived communities did not, suggesting that less wealthy people may be more likely to gamble in times of economic stress. When controlling for confounding variables the most deprived groups were 4.5 times as likely to experience a gambling-related argument or money issues than people living in the least deprived areas were.

### Other Factors

Studies that examined unique factors affecting harms include Jeffrey et al. (2019) who compared gamblers with their partners and found that gamblers were more likely to report individual harms that affect themselves. They also found that gamblers identified a wider range of harms and were better at identifying harms than their partners. This suggests that as well as experiencing more harms than affected others, gamblers are more aware of those harms and therefore may be more consciously impacted. Li et al. (2017) also found that harms in all areas accumulated more quickly in gamblers than in affected others.

Langham et al. (2017) found that an individuals’ sense of coherence correlated strongly with gambling harms. Sense of coherence is the extent to which someone feels confident in the predictability of his or her environment, and that things will generally turn out as expected.

Binde (2016) looked specifically at harms in the context of an individual’s place of work and found that signs of a problem gambler at work included excessive talk on gambling, gambling during breaks or during work time, borrowing money from colleagues, poor work performance, lateness or absence and more. Despite this, they did not compare different levels of employment or types of job role that could have provided a more thorough view of gambling harms and employment. Also, borrowing from colleagues should only be considered a gambling harm if the money is not paid back or the transaction causes a strain on the working relationship.

May-Chahal et al. (2017) investigated harms within the British prison population and found that although the prevalence of problem gambling in terms of the PGSI was higher in prisons, the prevalence of gambling behaviour prior to incarceration was significantly lower. They found that there was no link between PGSI score and criminal career, and no statistical link between gambling and drug use, however high rate offenders in their mid-20s were 5.3 times more likely to be frequent loss chasers than other categories. May-Chahal et al. (2017) also found that occasional gamblers were less likely to use alcohol or drugs in prison, with nearly 2/3 of the problem-gambling group abstaining completely from substance use. The researchers suggest that this may be because the individuals’ ‘addiction needs’ are being met by their gambling behaviour.

Three studies reported on how student status affected gambling harms, with Livazovic and Bojcic (2019) reporting that high-achieving students reported less psychological harms, and vocational students were significantly more at risk of harms than other students. However, there was only a weak correlation between success in school and harms from problem gambling. Melendez-Torres et al. (2019) also found that a reduced feeling of belonging at school was associated with more harms, but also higher rates of gambling, and Apinuntavech et al. (2012) found that average GPA was significantly lower in those who gambled, though only by a small margin. They also reported that students who had gambled were more likely to feel guilt, lie, experience depression, perceived poor health, and insomnia. With some students reporting substance use to manage stress, school absences and considering suicide.

Four studies also looked at how gambling harms present within a family unit, or how an individuals’ home life may influence their gambling. Anderson, Rempusheski, and Leedy (2018) examined senior family members and found that co-dependency within the family, where each person was expected to bail the other out, caused relationship breakdowns, stress and tension. Some participants expressed shame at spending their children’s trust funds or savings; however, participants also reported using the addiction model to neutralize shame and guilt. Ferrara, Franceschini, and Corsello (2018) and Larsen, Curtis, and Bjerregaard (2013) both found that an individuals’ home life affected their gambling habits. Ferrara, Franceschini, and Corsello (2018) stated that having separated parents increased the risk of gambling later in life, and Larsen, Curtis, and Bjerregaard (2013) found that the odds of showing one or more addictive behaviours increased in households without children. Perhaps suggesting that the presence of a traditional nuclear family unit is a protective factor against gambling harms. Alternatively, Livazovic and Bojcic (2019) found that family life and the parents’ level of education both had no significant effect on harms experienced, though they did affect the likelihood of taking part in risky behaviour.

## DISCUSSION

The results presented here suggest that there may be a health inequality in gambling harms, as several studies have found differences in the number and types of harms reported in different social groups. Although further analysis and investigation is necessary for a complete understanding of the distribution of gambling harms in society, the results suggest that there are differences that are dependent upon several factors. Studies such as Wardle et al. (2019), Castrén et al. (2017) and Tu, Gray, and Walton (2014) pose a particular concern as there are suggestions that certain groups experience more harms even when gambling less, presenting a health inequality that needs to be understood and addressed. In particular, several studies report differences between age groups, socioeconomic status, and gambling behaviour or play styles.

In applying the Standard Quality Assessment Criteria (Kmet, Lee, & Cook, 2004) we found that several studies were not robust in their quality control. In particular, studies scoring below 0.5 on the assessment may not be an accurate representation of gambling harms, whereas studies that scored above 0.95 may present the most reliable data on harm distribution (Table 3).

**Table 3.**
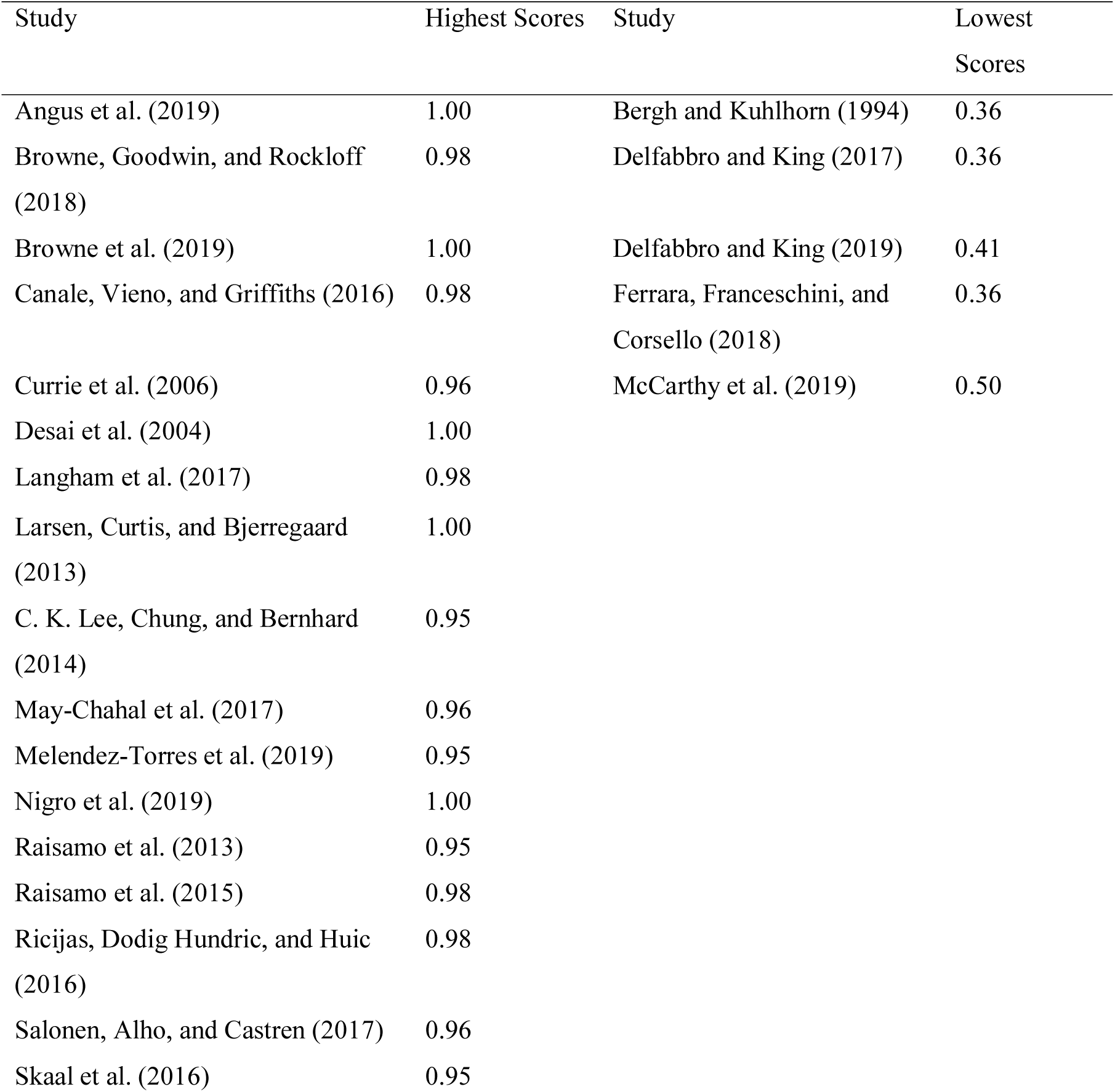
Highest and Lowest Quality Assessment Scores (Kmet, Lee, & Cook, 2004)

| Study | Highest Scores | Study | Lowest Scores |
| --- | --- | --- | --- |
| Angus et al. (2019) | 1.00 | Bergh and Kuhlhorn (1994) | 0.36 |
| Browne, Goodwin, and Rockloff (2018) | 0.98 | Delfabbro and King (2017) | 0.36 |
| Browne et al. (2019) | 1.00 | Delfabbro and King (2019) | 0.41 |
| Canale, Vieno, and Griffiths (2016) | 0.98 | Ferrara, Franceschini, and Corsello (2018) | 0.36 |
| Currie et al. (2006) | 0.96 | McCarthy et al. (2019) | 0.50 |
| Desai et al. (2004) | 1.00 |  |  |
| Langham et al. (2017) | 0.98 |  |  |

|  |  |
| --- | --- |
| Larsen, Curtis, and Bjerregaard<br>(2013) | 1.00 |
| C. K. Lee, Chung, and Bernhard<br>(2014) | 0.95 |
| May-Chahal et al. (2017) | 0.96 |
| Melendez-Torres et al. (2019) | 0.95 |
| Nigro et al. (2019) | 1.00 |
| Raisamo et al. (2013) | 0.95 |
| Raisamo et al. (2015) | 0.98 |
| Ricijas, Dodig Hundric, and Huic<br>(2016) | 0.98 |
| Salonen, Alho, and Castren (2017) | 0.96 |
| Skaal et al. (2016) | 0.95 |

One commonly reported factor in these high rated studies was the prevention paradox, and all studies that discuss this found evidence in favour of the paradox. The findings showed that the majority of harm impact is present in the lower PGSI rated groups (Angus et al., 2019; Browne & Rockloff, 2018; Canale, Vieno, & Griffiths, 2016; S. Raisamo et al., 2015), due to the larger population numbers in these categories. However, they also found that the highest severity of harm, such as alcohol use (Larsen, Curtis, & Bjerregaard, 2013) or severe psychological distress (Skaal et al., 2016), was most often present in the problem-gambling group. Angus et al. (2019) in particular reported significantly greater severity of harms in all domains for the clinical sample, even when controlling for those community participants who reported zero harms.

All but one of these high rated studies found no differences in harm presentation between genders. Where differences were reported males gambled more often than females with higher frequency of play and higher spends, and S. Raisamo et al. (2015) showed that when controlling for frequency of play gender differences were no longer significant. For example, Nigro et al. (2019) found that being male predicted level of risk for gambling, but also that higher involvement in gambling led to an increased risk of depression and memory impairment. The single outlier, Salonen, Alho, and Castrén (2017), found that occasional gambling increased in women 18-24 between the years 2011 and 2015, and only females felt more harms despite men 18-24 also increasing their gambling behaviour.

All but two studies (Browne, Goodwin, & Rockloff, 2018; Melendez-Torres et al., 2019) investigating age found significant differences across groups, with the majority of harms and dysfunctional symptomology reported in ages below 40 (Browne et al., 2019; Canale, Vieno, & Griffiths, 2016; Currie et al., 2006; Desai et al., 2004; Raisamo et al., 2013; S. Raisamo et al., 2015). Younger age groups were specifically linked to increased alcohol abuse, substance use, incarceration, bankruptcy and depression (Desai et al., 2004). In contrast Browne, Goodwin, and Rockloff (2018) found no differences, whereas Melendez-Torres et al. (2019) found that harms actually increased with age. Despite this they used a sample of school-aged children and adolescents and so the results cannot be compared to adult participants.

One highly rated study identified trait impulsivity as the most important risk factor in gambling, and suggested that impulsivity mediated the link between harms and age or gender (Browne et al., 2019). Sense of coherence also significantly correlated with gambling harms, with a weaker sense of coherence leading to increased harms. Along with impulsivity, sense of coherence could point to a personality type that increases the risk of harms from gambling. Moreover, evidence has shown that impulsivity is high in younger age groups, who appear to display more gambling harms than older individuals do.

Low household income was also found to be a significant predictor of gambling harms (Angus et al., 2019; Browne, Goodwin, & Rockloff, 2018; Browne et al., 2019; Currie et al., 2006). However, (Browne, Goodwin, & Rockloff, 2018) did report that the difference in harm risk between $15-30k AUD and $101-$150k was less than 5 points. (Browne et al., 2019) and (Skaal et al., 2016) also found that unemployment was related to increased harms, and (Skaal et al., 2016) reported that participants living in urban areas reported more psychological distress and alcohol problems than those from peri-urban areas. In contrast to these results (Melendez-Torres et al., 2019) reported more harms amongst the most affluent participants, however they also found that these individuals had higher frequency of play.

As previously stated those that play more frequently and spend more were found to have the highest number of harms (Canale, Vieno, & Griffiths, 2016; Nigro et al., 2019; Raisamo et al., 2013), and (Currie et al., 2006) found that harms increased significantly when an individual gambled more often than once per week. The choice of game was also found to be significant, with (Currie et al., 2006) reporting more harms in EGM users, instant win tickets, bingo and casinos, and (Ricijas, Dodig Hundric, & Huic, 2016) reporting more harms amongst sports bettors, VLT players and virtual bettors.

Two studies reported that non-white participants experienced more harms (Currie et al., 2006; Melendez-Torres et al., 2019), despite white individuals gambling less often. And three studies reported that lower education level (Browne et al., 2019; Nigro et al., 2019) and lower feelings of belonging within school (Melendez-Torres et al., 2019) lead to more gambling harms. (Ricijas, Dodig Hundric, & Huic, 2016) also found that superstitious beliefs were related to gambling harm, and (May-Chahal et al., 2017) reported that problem gambling as rated by the PGSI was higher among a prison population than the general population. Despite this they found that rates of problem gambling were lower among this group before incarceration, suggesting that the gambling problem developed within the prison system.

The final high rated study (C. K. Lee, Chung, & Bernhard, 2014) found that motivations of excitement, escape and challenge were linked to positive outcomes from gambling, whereas money as a motivator led to harmful consequences. Despite this another highly rated study (Browne et al., 2019) found that an individual’s reason to gamble, or motivation, had no impact on harms and so further research is needed to confirm or deny this association.

It was stated by Susana Jiménez Murcia that, “we need to use different treatments for each sub-group of pathological gamblers” (Plataforma SINC). Murcia is the co-author of a study that found that there are four distinct types of gambler (Álvarez-Moya et al., 2010). The team concluded that out of the four sub-types only one category of gambler suggested significant pathology, though all were compulsive with differing severity levels, comorbidity and personality profiles. Future research could investigate not only the distribution of harms across society, but also further understand these sub-types of gambler, attribute these to specific groups or personality profiles, and compare and validate the results against this previous work. This future research should also broaden the participant base as Álvarez-Moya et al. (2010) only investigated self-reporting slot machine gamblers, meaning their results may not be complete, or may not be generalizable.

To conclude, our review strongly suggests that the distribution of harms in the population is affected by a number of factors and presents some key signs to identify individuals who may be at risk. However, further research is needed to fully understand gambling harms and to confirm which individuals and groups are most at risk.

## LIMITATIONS

Despite these results it is important to consider the limitations of the study when reviewing the data. As was stated earlier, a large majority of the included papers investigated the general population of Australia, and so these findings may not be generalizable to a global sample. Many of the studies also included harmful consequences that are not universally accepted. For example chasing losses and betting above affordable means may be behaviours that lead to harms, and borrowing money could be considered a predictor of harms such as debt or relationship conflict.

## Data Availability

Data is publicly available on Web of Science and Scopus

## APPENDICES

Additional File 1.

The PRISMA Checklist

Table showing the PRISMA checklist completed in relation to this review as a PDF

Additional File 2.

Full Search Report

Full Electronic Report of Web of Science Search conducted 20^th^ February 2020 as a PDF

Additional File 3.

Table of Quality Checks

Table of the quality assessment completed by researchers as a PDF

Additional File 4.

Table of Extracted Data

Full table of data extracted from included studies as a PDF

## ACKNOWLEDGMENTS

We would like to thank Samantha Jordan for completing quality checks on the studies.

## DECLARATIONS

The authors declare no conflicts of interest in relation to this work

